# Comparative profiles of SARS-CoV-2 Spike-specific human milk antibodies elicited by mRNA- and adenovirus-based COVID-19 vaccines

**DOI:** 10.1101/2021.07.19.21260794

**Authors:** Xiaoqi Yang, Alisa Fox, Claire DeCarlo, Caroline Norris, Samantha Griffin, Sophie Wedekind, James M. Flanagan, Natalie Shenker, Rebecca L. Powell

## Abstract

Numerous COVID-19 vaccines are authorized globally. To date, ∼71% of doses are comprised of the Pfizer/BioNTech vaccine, and ∼17% the Moderna/NIH vaccine, both of which are mRNA-based. The chimpanzee Ad-based Oxford/AstraZeneca (AZ) vaccine comprises ∼9%, while the Johnson & Johnson/Janssen (J&J) human adenovirus (Ad26) vaccine ranks 4^th^ at ∼2% [1]. No COVID-19 vaccines are yet available for children 0-4. One method to protect this population may be passive immunization via antibodies (Abs) provided in the milk of a lactating vaccinated person. Our early work [2] and other reports [3-5] have demonstrated that unlike the post-SARS-CoV-2 infection milk Ab profile, which is rich in specific secretory (s)IgA, the vaccine response is highly IgG-dominant. In this report, we present a comparative assessment of the milk Ab response elicited by Pfizer, Moderna, J&J, and AZ vaccines. This analysis revealed 86% -100% of mRNA vaccine recipient milk exhibited Spike-specific IgG endpoint titers, which were 12 – 28-fold higher than those measured for Ad vaccine recipient milk. Ad-based vaccines elicited Spike-specific milk IgG in only 33%-38% of recipients. Specific IgA was measured in 52%-71% of mRNA vaccine recipient milk and 17%-23% of Ad vaccine recipient milk. J&J recipient milk exhibited significantly lower IgA than Moderna recipients, and AZ recipients exhibited significantly lower IgA titers than Moderna and Pfizer. <50% of milk of any group exhibited specific secretory Ab, with Moderna recipient IgA titers measuring significantly higher than AZ. Moderna appeared to most frequently elicit >2-fold increases in specific secretory Ab titer relative to pre-vaccine sample. These data indicate that current Ad-based COVID-19 vaccines poorly elicit Spike-specific Ab in milk compared to mRNA-based vaccines and that mRNA vaccines are preferred for immunizing the lactating population. This study highlights the need to design vaccines better aimed at eliciting an optimal milk Ab response.

## Background

SARS-CoV-2, the causative virus of the COVID-19 pandemic, has infected >340 million people worldwide, causing >5.6 million deaths. Though pediatric COVID-19 is mild in most cases, ∼10% of infected infants <1 year old experience illness requiring advanced care and have the highest ICU admission and case-fatality rates globally of all children (∼1.41% and ∼0.58%, respectively) [6-9]. The need for advanced care for survival in even a small minority of SARS-CoV-2-infected children is highly relevant in terms of the impact of pediatric infection in low-income settings where such care may not be available. Critically, a recent meta-analysis found that COVID-19 pediatric case-fatality rates (CFR) were significantly higher in low and middle-income countries (LMIC) compared to high-income countries (HIC) (0.24% compared to 0.01%), with the highest CFR in infants <1 year old (0.07% in HIC and 1.30% in LMIC) [9]. Notably, even asymptomatic infection can lead to ‘ Multisystem Inflammatory Syndrome in Children’ (MIS-C), a rare but potentially deadly inflammatory condition and young children are likely responsible for a significant amount of SARS-CoV-2 dissemination [10, 11] [12-15]. Clearly, protecting this population from infection remains essential, especially as variants emerge that are more transmissible or possibly more pathogenic in children [13-15]. Indeed, a recent study found that children are in fact at similar risk of SARS-CoV-2 infection as adults [16], and during the Delta (B.1.617.2) wave of infections in the USA, new cases in children aged 0-4 surpassed those among adults 65+ at times, an epidemiological feature not previously observed during the pandemic [17]. Fortunately, Numerous COVID-19 vaccine candidates employing a variety of novel and conventional platforms have been authorized globally. To date, ∼71% of the world’s doses have been comprised of the Pfizer/BioNTech vaccine, and ∼17% the Moderna/NIH vaccine, both of which are mRNA-based. The chimpanzee Ad-based Oxford/AstraZeneca (AZ) vaccine comprises ∼9% of global doses, while the Johnson & Johnson/Janssen (J&J) human adenovirus (Ad26) vaccine ranks 4^th^ at ∼2% [1]. Importantly, none of these vaccines are yet authorized for use in infants or young children, and no COVID-19 vaccine candidates appear to be under investigation for use in infants < 6 months. Although this is in part due to the potential for these infants to have received vaccine- or infection-induced maternal IgG in utero, which could interfere with infant vaccine efficacy, optimal placental IgG transfer to these infants is far from guaranteed, as it requires timely maternal vaccine access, as well as surmounting the significant vaccine hesitancy among pregnant people [18, 19]. Importantly, maternal antibodies (Abs) wane from birth until they are undetectable by 6-12□months, with the kinetics of this decline being highly dependent on the initial titers transferred [20]. As such, the passive immunity of the Abs provided through human milk-feeding by a COVID-19-vaccinated mother or milk donor is a critical mechanism to protect young children from SARS-CoV-2 infection and pathology as the pandemic continues. Even for infants receiving high titers of maternal IgG in utero, milk Ab can provide critical complementary mucosal protection [21, 22]. Particularly in LMIC, vaccine access for young children is not likely to be rapid or straightforward, and children in these settings are more likely to breastfeed for 2 years or more; therefore, vaccine Ab transferred via milk for all breastfeeding children remains especially relevant. It is therefore of utmost importance to methodically assess the human milk immune response to each of the novel COVID-19 vaccines.

Mature human milk contains ∼0.6mg/mL total Ab, though there is great variation among women sampled [23]. Milk IgG originates predominantly from serum with some local production in specific cases, though generally IgG comprises only ∼2% of total milk Ab [24]. Approximately 90% of total milk Ab is IgA and ∼8% IgM, nearly all in secretory (s) form (sIgA/sIgM; polymeric Abs complexed to j-chain and secretory component (SC) proteins) [24-26]. Nearly all sIgA/sIgM derives from the gut-associated lymphoid tissue (GALT), via the *entero-mammary link*, though there is also homing of B cells from other mucosal-associated lymphoid tissue (MALT), i.e. the respiratory system to the mammary gland. The SC protein is a cleaved segment of the polymeric immunoglobulin receptor (pIgR) which transports this GALT/MALT-derived Abs into the milk.

Relatively few comprehensive studies exist examining the Ab response in milk after vaccination. The few studies that have examined the milk Ab response after influenza, pertussis, meningococcal and pneumococcal vaccination have generally found specific IgG and/or IgA that tends to mirror the serum Ab response, though none of these studies measured secretory Ab or determined if sIgA was elicited, and data regarding the protective capacity of these milk Abs is conflicting or confounded by the effects of placentally-transferred Ab [22, 27-33]. Determining whether or not secretory Abs are elicited in milk after infection or vaccination is critical, as this Ab class is highly stable and resistant to enzymatic degradation in all mucosae - not only in the infant oral/nasal cavity, but in the airways and GI tract as well [24, 34].

Our comprehensive study to assess the SARS-CoV-2-specific Ab response in human milk after infection has so far determined that SARS-CoV-2 infection elicits a robust specific milk IgA response in ∼90% of cases, which is very strongly correlated with a robust specific secretory Ab response [35]. This sIgA response is highly durable over time, neutralizing, and dominant compared to the specific milk IgG response in terms of potency and number of responders [35, 36]. Our early analysis of the milk Ab response to mRNA-based COVID-19 vaccination found Spike-specific IgG to be dominant in milk compared to IgA 14 days after the 2^nd^ vaccine dose, with all samples containing significant levels of specific IgG, with 80% of samples exhibiting high IgG endpoint titers. Conversely, only 50% of post-vaccine milk samples contained Spike-specific secretory Ab, which were of low-titer. Since this report, other groups have confirmed the IgG-dominant milk Ab profile after mRNA-based vaccination, noting that the observed high titers are induced after the 2^nd^, boosting dose of vaccine [3-5]. It has also been observed that specific milk IgA responses are elicited by the initial, priming dose of mRNA vaccine, but poorly boosted after the 2^nd^ dose [4]. mRNA-based vaccination has also been shown to elicit neutralizing activity in milk [3]. No further studies since our early report have assessed the specific secretory Ab response to COVID-19 vaccination, and no studies to date have reported on the milk Ab profile induced by the J&J vaccine or a complete regimen (2 doses) of the AZ vaccine [37].

Herein we describe the vaccine-elicited, SARS-CoV-2 Spike-specific Ab profile of pairs of milk samples obtained from 54 individual donors within 1 week before vaccination and 14 (Moderna/Pfizer/AZ) or 21-35 days (J&J) after completion of a COVID-19 vaccine regimen. Samples were assayed for specific IgA, IgG, and secretory Ab against the full trimeric SARS-CoV-2 Spike protein.

## Methods

### Study participants

Individuals were eligible to have their milk samples included in this analysis if they were lactating, had no history of a suspected or confirmed SARS-CoV-2 infection, and were scheduled to be or had recently been vaccinated with the Pfizer, Moderna, J&J, or AZ COVID-19 vaccine. If pre-vaccine milk samples were determined to be positive for SARS-CoV-2 IgA, samples were excluded from the present analysis. Pfizer, Moderna, and J&J sample collection was performed in the USA and approved by the Institutional Review Board (IRB) at Mount Sinai Hospital (IRB 19-01243). AZ recipient milk samples were collected from women living in England as part of the Breastmilk Epigenetics Cohort Study (WREC 15/WA/0111, IRAS 165531). All women gave informed consent after reading the related information sheets. Once consented into the study, participants were asked to collect approximately 30mL of milk per sample into a clean container using electronic or manual pumps. Milk was frozen in participants’ home freezer until samples were picked up and stored at -80°C until Ab testing.

### ELISA

Levels of SARS-CoV-2 Abs in human milk were measured as previously described [35]. Briefly, before Ab testing, milk samples were thawed, centrifuged at 800g for 15 min at room temperature, fat was removed, and supernatant transferred to a new tube. Centrifugation was repeated 2x to ensure removal of all cells and fat. Skimmed acellular milk was aliquoted and frozen at -80°C until testing. Milk was tested in separate assays measuring IgA, IgG, and secretory-type Ab reactivity (the secondary Ab used in this assay is specific for free and bound human secretory component). Half-area 96-well plates were coated with the full trimeric spike protein produced recombinantly as described [38]. Plates were incubated at 4°C overnight, washed in 0.1% Tween 20/PBS (PBS-T), and blocked in PBS-T/3% goat serum/0.5% milk powder for 1h at room temperature. Milk was used undiluted or titrated 4-fold to 1/4096 in 1% bovine serum albumin (BSA)/PBS and added to the plate. After 2h incubation at room temperature, plates were washed and incubated for 1h at room temperature with horseradish peroxidase-conjugated goat anti-human-IgA, goat anti-human-IgG (Fisher), or goat anti-human-secretory component (MuBio) diluted in 1% BSA/PBS. Plates were developed with 3,3’,5,5’-Tetramethylbenzidine (TMB) reagent followed by 2N hydrochloric acid (HCl) and read at 450nm on a BioTek Powerwave HT plate reader. Assays were performed in duplicate for each dilution and repeated 2x.

### Analytical Methods

Milk was defined as positive for Spike Ab if OD values measured using undiluted milk from COVID-19-recovered donors were more than two standard deviations (SD) above the mean ODs obtained from pre-vaccine samples. Endpoint dilution titers were determined from log-transformed titration curves using 4-parameter non-linear regression and an OD cutoff value of 1.0. Endpoint dilution positive cutoff values were determined as above. Mann-Whitney U tests were used to assess significant differences. Correlation analyses were performed using Spearman correlations. All statistical tests were performed in GraphPad Prism, were 2-tailed, and significance level was set at p-values < 0.05.

## Results

Fifty-four pairs of milk samples were obtained from vaccine recipients within 1 week before vaccination and 14 days (Pfizer/Moderna/AZ) or 21-35 days (J&J) after completion of the vaccine regimen. Twenty-one participants had received Pfizer vaccine, 14 had received Moderna vaccine, 13 had received J&J vaccine and 6 had received AZ vaccine. Milk was used in separate ELISAs measuring IgG, IgA, and secretory Ab binding against recombinant trimeric SARS-CoV-2 Spike [38]. As shown in Fig. 1, OD values were recorded at each 4-fold milk dilution assayed (undiluted to 1/4096 dilution). Titrated milk ODs were used to determine endpoint binding titers. It was found that 100% and 86% of Moderna and Pfizer recipient post-vaccine milk samples, respectively, contained positive levels of Spike-specific IgG, with endpoint titers surpassing the absolute positive cutoff value (mean endpoint titer = 120 and 190, respectively; Fig. 1a). The mean IgG titers of these mRNA vaccine groups were not significantly different. In contrast, both groups exhibited significantly higher specific milk IgG than milk obtained from J&J and AZ vaccine recipients, with only 38% and 33%, respectively, of these samples containing positive levels of specific IgG (mean endpoint titers = 10 and 6.6, respectively; p=0.0011 - <0.0001; Fig. 1a, 3^rd^ panel). It was further determined that 71% and 52% of Moderna and Pfizer recipient post-vaccine milk samples, respectively, contained positive levels of Spike-specific IgA, exhibiting similar mean endpoint titers of 19 and 24, respectively (Fig. 1b). Twenty-three percent of J&J and 17% of AZ vaccine recipient milk samples contained positive levels of Spike-specific IgA, exhibiting a mean endpoint titers of 15 and 5.6, which were significantly lower than that of the Moderna vaccine group (0.020<p<0.025; Fig. 1b, 3^rd^ panel). As well, the AZ group mean titer was significantly lower than the Pfizer group mean titer (p=0.030). Finally, it was determined that 43% and 48% of the Moderna and Pfizer recipient post-vaccine milk samples, respectively, contained positive levels of Spike-specific secretory Ab, exhibiting similar mean endpoint titers of 12 and 14, respectively (Fig. 1c). These endpoint titers were not significantly different from that of the J&J vaccine recipient milk samples (mean endpoint titer=8), 31% of which contained positive levels of Spike-specific secretory Ab, (Fig. 1c, 3^rd^ panel). However, the Moderna group mean titer was significantly higher than that of the AZ group (endpoint titer=2.7; p=0.0063), which did not include any samples exhibiting Spike-specific secretory Ab.

**Figure 1.**
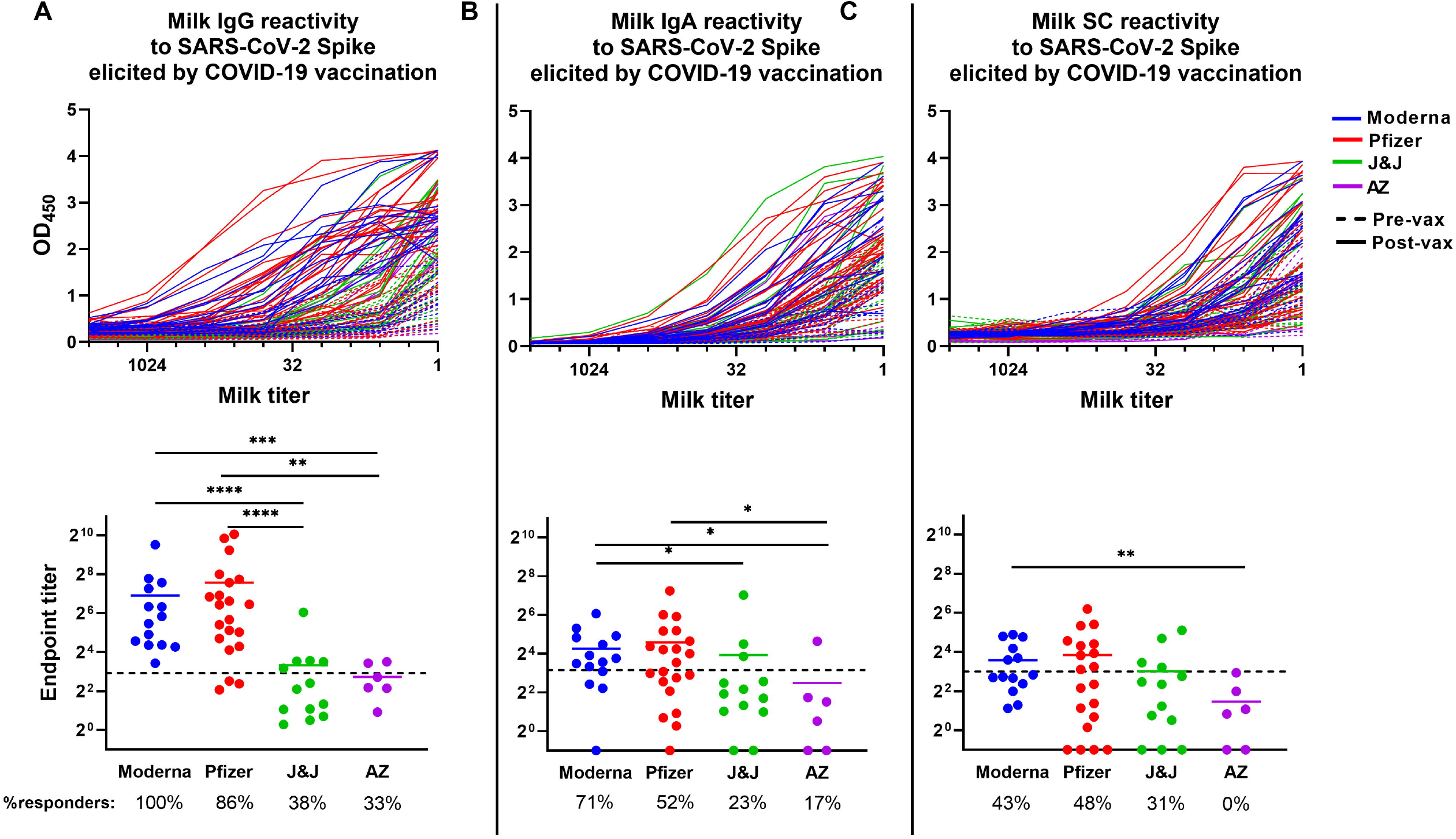
The COVID-19 vaccine-induced SARS-CoV-2 Spike-specific milk antibody response. Plates were coated with full-length recombinant trimeric SARS-CoV-2 Spike. Milk was titrated and added to the plate. Plates were washed and blocked and incubated with appropriate secondary antibody. Titration curves were plotted and used to calculated endpoint titers. (A) IgG titration curves (top panel) and endpoint titers (bottom panel). (B) IgA titration curves (top panel) and endpoint titers (bottom panel). (C) Secretory antibody titration curves (top panel) and endpoint titers (bottom panel). Dotted lines indicate positive cutoff values. Responders (%): Percent of participants receiving this vaccine type exhibiting positive endpoint titers. ****p<0.0001; *** 0.0001<p<0.009; *0.009<p<0.01; *0.01<p<0.05. Mean values are shown in scatter plots.

Given the availability of pre-vaccine samples for all participants included in this study, and the stringent nature of our positive cutoff values that are a reflection of the natural human variation of ‘background’ Ab activity of SARS-CoV-2-naïve milk, we next sought to compare individual samples pre- and post-vaccine ELISA data, designating >2-fold increase in endpoint titer as a significant change in Ab reactivity on an individual basis (Fig 2). This analysis determined that 100% of Moderna, Pfizer, and AZ recipient milk samples exhibited significant relative increases in IgG reactivity against Spike after vaccination, while only 62% of J&J recipient milk samples exhibited this effect (Fig. 2a). In terms of IgA reactivity, 93% of Moderna recipient milk samples exhibited a notable increase, compared to 75% of Pfizer recipients, 54% of J&J recipients, and 50% of AZ recipients (Fig. 2b). Finally, examining the secretory Ab data, 86% of Moderna recipient milk samples exhibited > 2-fold increase in endpoint titer, while 55% of Pfizer, 62% of J&J, and 50% of AZ recipient milk samples exhibited such an increase (Fig. 2c). Using these data, the mean relative fold increase in Ab reactivity from pre-vaccine to post-vaccine was compared among the vaccine groups. It was found that Moderna and Pfizer recipients exhibited similar increases in Spike-specific milk IgG reactivity (mean endpoint titer increases = 90x and 126x, respectively), while J&J and AZ recipients exhibited significantly lower relative IgG titer increases (mean=12x and 8x, respectively; p<0.0001; Fig. 2d). J&J recipients exhibited significantly greater relative IgG increases compared to AZ (p=0.0365). In terms of the IgA response, Moderna vaccine recipients exhibited significantly greater relative increases compared to Pfizer vaccine recipients (mean endpoint titer increase=23x versus 6x; p=0.04; Fig. 2d). Moderna group increases were also significantly higher compared to J&J recipient milk samples, which exhibited a mean relative endpoint titer increase of 4x (p=0.003), and AZ recipient milk samples (mean endpoint increase=3x; p=0.0095). All 4 groups exhibited similarly small relative increases in Spike-specific secretory Ab reactivity post-vaccine (mean endpoint titer increase = 5x - 7x; Fig 2d).

**Figure 2.**
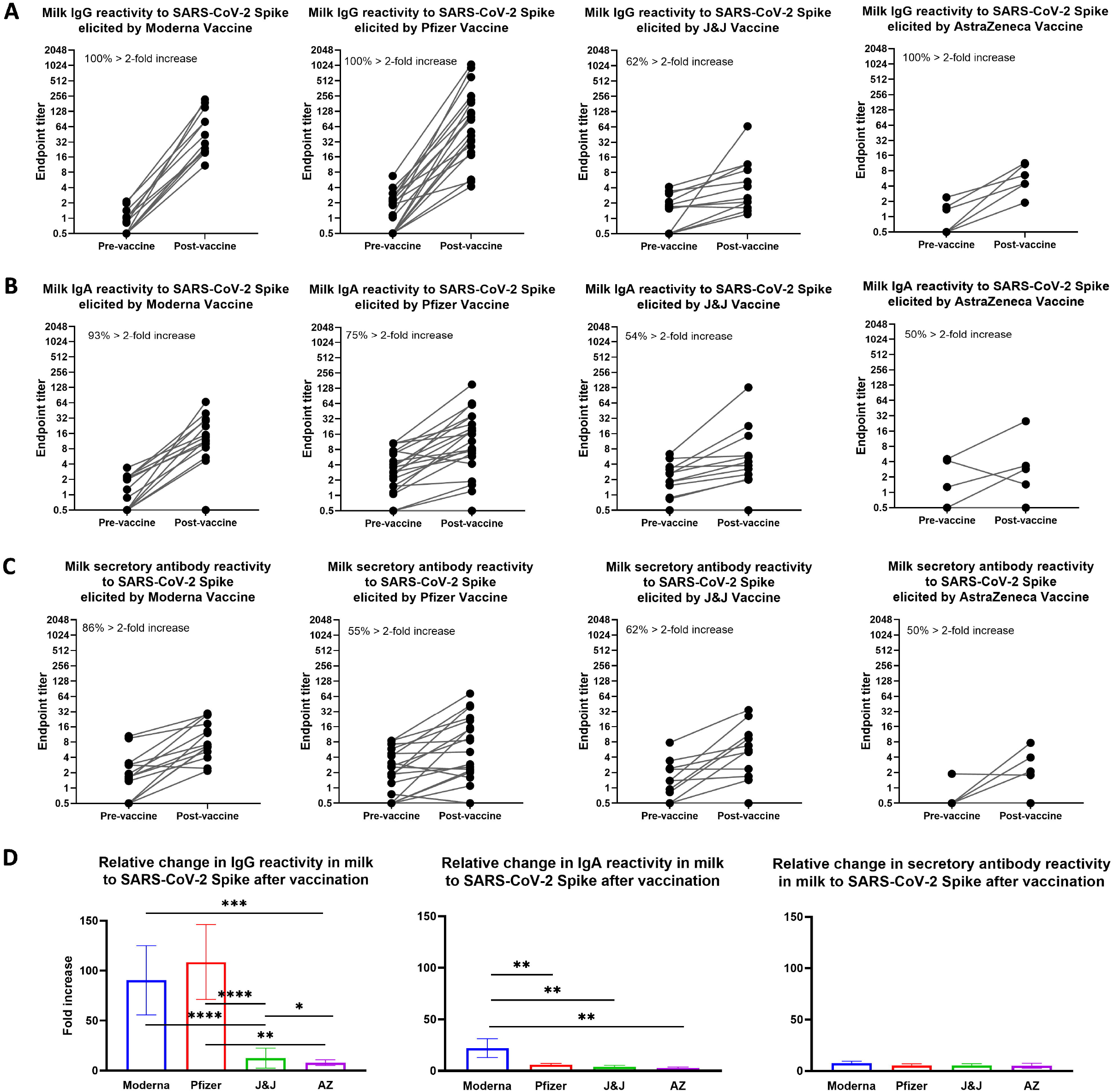
Relative increases in milk antibody reactivity to SARS-CoV-2 Spike elicited by COVID-19 vaccination. Endpoint titers were determined as in Fig. 1 for pairs of milk samples obtained from participants within 1 week before and 14 (Pfizer/Moderna/AZ) or 21-35 (J&J) days after completion of a vaccine regimen. Pre-vaccine and post-vaccine endpoint titers are shown for: (A) IgG reactivity. (B) IgA reactivity. (C) Secretory antibody reactivity. 1^st^ panel, Moderna vaccine; 2^nd^ panel, Pfizer vaccine; 3^rd^ panel, J&J vaccine; 4^th^ AZ vaccine. Each plot indicates the percent of vaccine recipients exhibiting > 2-fold increase in specific endpoint titer between pre- and post-vaccine milk samples. (D) Fold increases in endpoint titers are shown for each vaccine group as indicated.. ****p<0.0001; *** 0.0001<p<0.009; *0.009<p<0.01; *0.01<p<0.05. Mean values with SEM are shown.

ELISA endpoint binding titers for each assay were compared in separate Spearman correlation analyses for each vaccine group (IgG v IgA; IgA v SC; IgG v SC). Moderna recipient milk IgG and IgA titers were not correlated, while Pfizer recipient milk IgG and IgA titers were shown to exhibit some positive correlation (r=0.47; p=0.024; Fig. 3a, 2^nd^ panel). In contrast, J&J recipient milk IgG and IgA titers were shown to exhibit a moderately strong positive correlation (r=0.76; p=0.0033; Fig. 3a, 3^rd^ panel). AZ recipient samples did not exhibit correlated IgG and IgA titers, the sample number was low for this group. Notably, Moderna, Pfizer, and J&J recipient milk IgA and secretory Ab titers were found to exhibit moderately strong positive correlation (r=0.71, 0.66, and 0.78; p=0.0058, 0.0010, 0.0064, respectively; Fig. 3b). AZ samples, despite the low sample number, exhibited very strongly positive correlation for IgA and secretory Ab titers (r=0.94; p=0.0167). Similar to the IgG v IgA data, Moderna recipient milk IgG and secretory Ab titers were not correlated, Pfizer recipient titers were somewhat positively correlated (r=0.46; p=0.033; Fig. 3c, 2^nd^ panel), and J&J recipient milk IgG and secretory Ab titers were shown to exhibit a moderately strong positive correlation (r=0.70; p=0.0097), and AZ data was not correlated.

**Figure 3.**
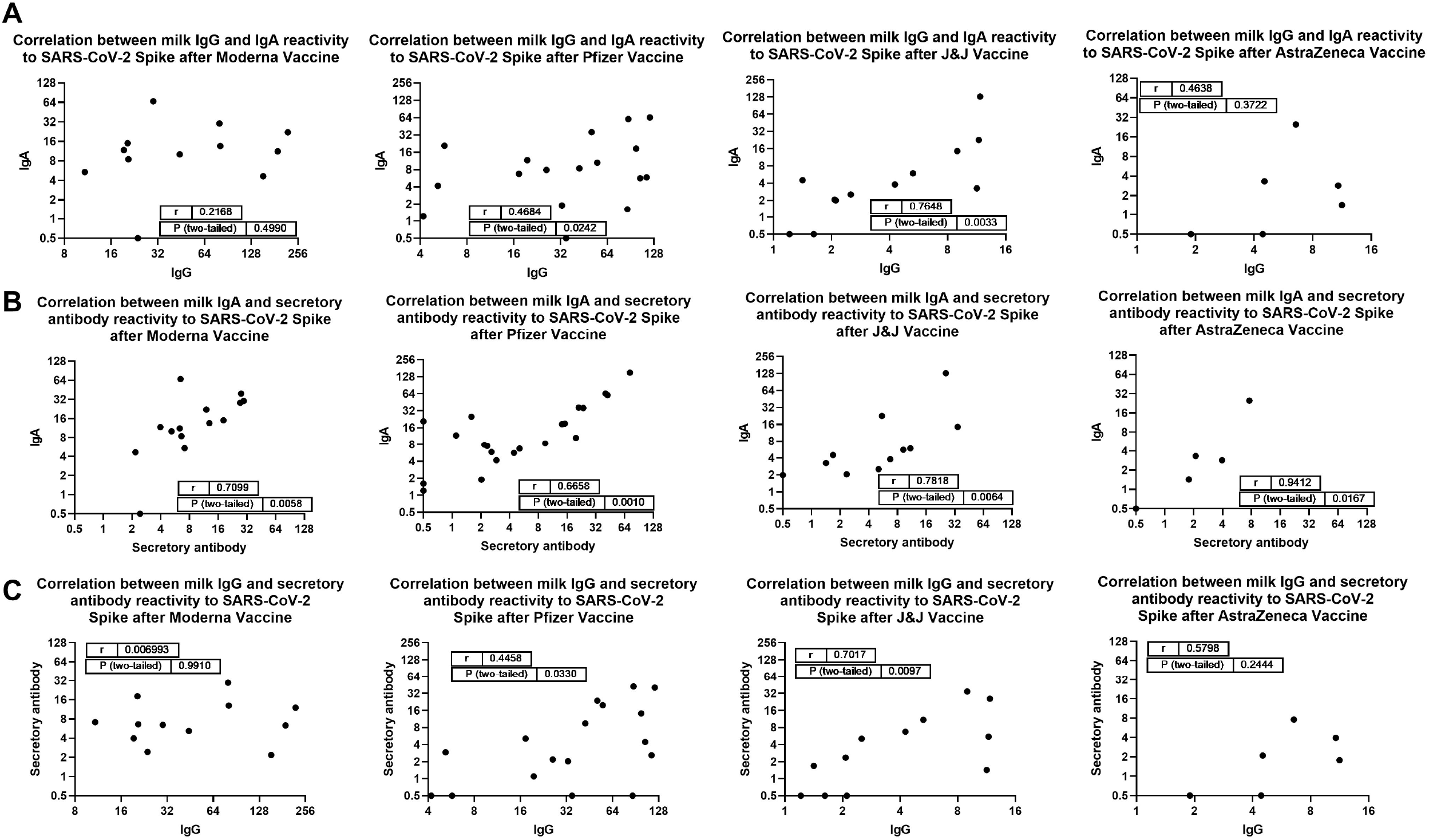
Correlation analyses. Endpoint titers calculated in Fig.1 were used in 2-tailed Spearman correlation tests. (A) Correlation analysis of IgA versus IgG reactivity. (B) Correlation analysis of IgA versus secretory antibody reactivity. (C) Correlation analysis of IgG versus secretory antibody reactivity. 1^st^ panel, Moderna vaccine; 2^nd^ panel, Pfizer vaccine; 3^rd^ panel, J&J vaccine; 4^th^ panel, AZ vaccine.

## Discussion

Though certainly the priority is to vaccinate all people as quickly as possible, the availability of multiple COVID-19 vaccines has raised the question globally, particularly the more vaccine-hesitant, as to which vaccine is best [39]. There is currently a lack of pertinent data, particularly for specific populations such as those who are lactating, that might be used to make an informed decision. The Pfizer and Moderna COVID-19 vaccines are both 2-dose regimens, intended to be delivered 3 and 4 weeks apart, respectively, and both consist of lipid-encapsulated, full length Spike mRNA, both incorporating virtually identical trinucleotide cap analogs, optimized Spike sequences, and N1-methylpseudouridine; however, their lipid carriers differ and the Pfizer vaccine consists of 30ug of RNA while Moderna includes 100ug [40]. The J&J vaccine is a recombinant, replication-incompetent Ad26 vector encoding full-length SARS-CoV-2 spike, authorized as a single dose regimen. The AZ vaccine also utilizes an adenoviral-vectored platform that is non-replicating and facilitates cellular expression of the SARS-CoV-2 Spike, though AZ vaccine uses a chimp adenovirus and was authorized as a 2-dose regimen [40]. For these Ad-based vaccines, vector-mediated innate immune responses, anti-vector and Spike-directed immunogenicity would be expected to differ, with an unknown impact of the generation of milk Abs. Notably, J&J and AZ vaccines have less stringent cold-chain requirements, and therefore may be more accessible to certain rural or otherwise vulnerable populations. The single-dose formulation of J&J also may suit such contexts.

Published immunogenicity data of these vaccines, though not assessed together in a single study, demonstrated that all 4 vaccines elicited positive specific (Wuhan strain) serum IgG titers in 100% of participants, with titers ∼10-20x higher for the mRNA vaccines compared to the Ad-based vaccines; however, whether neutralization potencies are significantly different among the vaccine types is not clear [41-44]. Serum IgA titers were not measured as part of clinical trial data, though a few recent reports have described the vaccine-induced serum IgA response to the mRNA vaccines, which appear moderate after the 1^st^ dose and do not appear to be significantly boosted after dose 2 [3, 4]. This present analysis of the milk Ab response to COVID-19 vaccination has furthered our initial finding that Spike-specific IgG is dominantly elicited by 14 days after the 2^nd^ dose of an mRNA-based vaccine, and that both Moderna and Pfizer perform equally in this regard. Importantly, these data demonstrate that Ad-based COVID-19 vaccines elicit far less IgG in milk, with far fewer vaccinees exhibiting positive levels of specific milk IgG or notable increases in reactivity compared to baseline levels pre-vaccine. Though specific milk IgA production is only moderate for all vaccine types, confirming our previous preliminary mRNA vaccine data and that of others, Moderna recipient milk appears to contain the highest levels of IgA, which is considerably more pronounced when examining individual relative increases compared to pre-vaccine samples. The increased specific IgA production by Moderna recipients, which may be due to differential class switching to IgA after the longer 28 day interval between doses compared to Pfizer vaccine, has been noted previously in serum, and our present data demonstrates the effect is similar in milk [3]. Again, J&J and AZ recipient data suggest that in terms of mean milk IgA level and/or percent of responders, this vaccine type does not perform well relative to mRNA vaccines. Finally, measuring the critically important secretory Ab response, levels are low and responders are few for all vaccine types studied, though the Moderna group included 25% more responders with notable increases in secretory Ab activity compared to baseline levels.

Generally, our IgG versus IgA correlation analysis of the mRNA vaccine data found these titers were at best only mildly related, with Moderna recipients exhibiting no apparent relationship; notably, these data were nearly identical for the IgG versus secretory Ab analysis. This finding is likely in part a reflection of the strong IgG and poor IgA boosting by the 2^nd^ mRNA vaccine dose noted previously [4]. In contrast, J&J recipient milk samples exhibited a relatively strong correlation between IgG and IgA reactivities, suggesting that if a measurable milk Ab response is generated by the J&J vaccine in a given individual, it is likely to exhibit balanced IgG/IgA class-switching. AZ recipients did not exhibit this correlation, possibly due to the small sample number in this group, or for similar reasons as the mRNA vaccine recipients. In terms of our analysis of the relationship between vaccine-induced IgA and secretory Ab, which is a proxy measure for assessing the potency of the sIgA response, the increased number of samples compared to our early report facilitated the determination that there was a moderately strong correlation between IgA and secretory Ab elicited by the mRNA vaccines [2]. This suggests that much of the IgA measured in milk is sIgA; however, compared to our post-infection data, the correlation is considerably weaker, suggesting that considerably more vaccine-induced monomeric IgA, like IgG, is entering the milk via passive transfer from serum, with mucosal-origin plasma cells producing dimeric IgA also transiting to the mammary gland to secrete sIgA into the milk [35]. J&J recipient milk samples demonstrated a somewhat stronger correlation between IgA and secretory Ab titers, possibly indicating that despite the low levels and number of responders exhibiting an IgA or secretory Ab response, if these responses in milk are measurable, they are more likely to be mucosally-derived sIgA, with less serum-derived monomeric IgA entering the milk compared to mRNA vaccines. AZ recipient milk samples, despite the very small sample number, exhibited very high correlation of IgA and secretory Ab titers, suggesting that if the titers in milk overall elicited by this vaccine can be improved, it may promote the most ideal milk Ab response. Given the small sample size studied here, more study is needed.

Intramuscular (IM) vaccines have been shown previously to generate mucosal Ab, including Abs in milk, though whether IM vaccination tends to elicit secretory Abs, which would be expected to be the most protective class in a mucosal environment, has generally not been addressed [22, 27-33]. Though secretory Abs are vital to the milk Ab defense system, monomeric, non-secretory Abs, originating from serum as well as locally, likely also contribute to the milk Ab defense system, particularly in the case of vaccination [45]. The data presented herein indicate that J&J and AZ Ad-based vaccines poorly elicit Spike-specific Ab in milk compared to mRNA-based vaccines, and suggest that Moderna vaccine elicits a superior milk (s)IgA response, though secretory Ab titers were fairly low for all vaccine types studied. In areas where COVID-19 vaccines are freely available, in terms of potential passive immunization of infants and young children via breastfeeding or milk sharing, these data suggest that Moderna vaccine may be the most ideal, while Ad-based vaccines are not.

## Data Availability

All data is provided in the manuscript, and raw data is available upon request.

## Acknowledgements

We are indebted to our milk donors. Spike protein was gifted by the Krammer Lab. This work was supported by the Icahn School of Medicine at Mount Sinai (RP). The BECS study was initially funded by Breast Cancer Now (JF) and currently by UKRI Future Leaders Fellowship (NS). The authors acknowledge infrastructure support from the Imperial Experimental Cancer Medicine Centre, Cancer Research UK Imperial Centre, the National Institute for Health Research Imperial Biomedical Research Center and the Ovarian Cancer Action Research Centre.

